# Serial population-based sero-surveys for COVID-19 in low and high transmission neighborhoods of urban Pakistan

**DOI:** 10.1101/2020.07.28.20163451

**Authors:** Muhammad Imran Nisar, Nadia Ansari, Farah Khalid, Mashal Amin, Hamna Shahbaz, Aneeta Hotwani, Najeeb Rehman, Sierra Pugh, Arjumand Rizvi, Arslan Memon, Zahoor Ahmed, Ashfaque Ahmed, Junaid Iqbal, Ali Faisal Saleem, Uzma Bashir Aamir, Daniel B Larremore, Bailey Fosdick, Fyezah Jehan

## Abstract

**Objective:** To determine population-based estimates of COVID-19 in a densely populated urban community of Karachi, Pakistan.

**Methods:** Three cross-sectional surveys were conducted in April, June and August in low- and high-transmission neighborhoods of Karachi. Participants were randomly selected to provide blood for Elecsys® immunoassay for detection of anti-SARS-CoV-2 antibodies. Bayesian regression model was used to estimate seroprevalence after adjusting for the demographic characteristics of each district. Conditional Risk of Infection (CRI) with 95% confidence interval was calculated using a non-parametric bootstrap of households. Infection fatality rates (IFR) were estimated.

**Results:** We enrolled 3005 participants from 623 households. In Phase 2, the adjusted seroprevalence was estimated as 8.7% (95% CI 5.1-13.1) and 15.1% (95% CI 9.4 -21.7) in low and high transmission areas respectively, compared to 0.2% (95% CI 0-0.7) and 0.4% (95% CI 0 - 1.3) in Phase 1. In Phase 3, it was 12.8% (95% CI 8.3 – 17.7) and 21.5% (95% CI 15.6-28) in low and high transmission areas, respectively. CRI was 0.31 (95% CI 0.16-0.47) and 0.41(95% CI 0.28-0.52) in low and high transmission neighborhoods respectively in Phase 2. Similar trends were observed in Phase 3. Only 5.4% of participants who tested positive for COVID-19 were symptomatic. IFR was 1.66% in phase 1, 0.37% in Phase 2 and 0.26% in Phase 3.

**Conclusion:** Initial rapid increase in seroprevalence was followed by a plateau in the later phase of the pandemic in Karachi. Continuing rounds of seroprevalence studies have the potential to fully characterize the pandemic in this geography.

## INTRODUCTION

The COVID-19 pandemic has resulted in more than 62 million confirmed cases and over 1.4 million deaths globally, case fatality rate (CFR) around 5.4% and infection fatality rate (IFR) of 0.9% (1, 2). As the world hastened to respond to the global health crisis, the pandemic revealed numerous cracks in health-care systems (3). Pakistan was amongst the first low- and middle-income countries (LMICs) to be affected by the pandemic and has reported 398,024 cases and 8,025 deaths (CFR 2.51 %) (1, 4). The CFR has been variable; as high as 5.11% (KPK) and as low as 0.71% (Gilgit Baltistan) with 1.82%, 1.80% and 1. 28% reported for Punjab, Sindh and Baluchistan respectively (5).

The demographic characteristics of Pakistan are typical of most LMICs with 41% of the population under the age of 15 (6). The societal construct in Pakistan boasts a patrilineal joint family system, intergenerational co-residence of family members and average household sizes of 6 or greater (7). This is important because recent evidence indicates that households with individuals above 60 years of age are at risk of life-threatening manifestations of the disease. (8)., Karachi, is home to people from varying diasporas across Pakistan, and the epicenter of the epidemic since 26^th^ Feb 2020, reporting the largest number of cases in Pakistan (84,232) and accounting for 28% of all COVID-19 cases in the country (9). Crowded neighborhoods and urban slum dwellings along with poor adherence to mitigation measures in the city may have expeditiously increased infection transmission (10).

Surveillance systems form the basis for tracking COVID-19 cases, testing populations at risk and carrying out contact tracing, thereby, representing the key components of the public health response (11). Facility-based surveillance efforts are likely to miss mild and asymptomatic cases, as bolstered by evidence from a WHO-China Joint Commission Report and several published studies which indicate that between 5-80% of sero-positive patients are noted to be asymptomatic. Therefore, household targeted serological testing can decrease biases arising from selective testing and generate concrete evidence on the role that asymptomatic transmission of infection plays in rapidly increasing infection rates (12). Household transmission is of special concern in congested neighborhoods of metropolitan cities when lockdown measures are in place. Secondary transmission from index cases in households using prospective follow-up and active symptom monitoring with nasopharyngeal polymerase chain reaction (NP-PCR) has indicated household attack rates as high as 32.4% (95% CI 22.4%–44.4%) (13). However, this exercise is resource intensive and transmission may differ between symptomatic and asymptomatic households, as symptomatic individuals are more likely to transmit the virus (14-17).

Using seroprevalence data, the conditional risk of infection (CRI), namely, the probability that an individual in a household is infected given that another household member is infected, can serve as a related index of infection within household (18).

With this approach, we estimated changes in seroprevalence in low- and high-transmission neighborhoods of Karachi between April and August 2020 through cross-sectional sero-surveys by adapting the World Health Organization (WHO) Unity protocol. We then compared the seroprevalence to total reported positive tests in the area. This was done to understand the adequacy of reporting from NP-PCR testing and to determine the infection fatality rate. We also determined age - gender stratified estimates of seroprevalence and assessed the conditional risk of infection (CRI) (19).

## MATERIALS and METHODS

### Study Participants and sample collection

We conducted the study in two areas. Four sub-administrative units (union councils) of District East were selected as high-transmission areas based on the number of cases reported by the provincial government. (Supplementary Figure 1) The study was approved by the Aga Khan University Ethical Review Committee (AKU ERC).

Three cross-sectional surveys were sequentially performed at the household level, between April 15-25, June 25-July 11 and August 17-22, 2020. Four research teams, each comprising of one data collector and one phlebotomist collected data and serology samples. A detailed line-list of cases was available in District East which allowed households to be selected through systematic random sampling as follows: A case was randomly identified from the line listing that served as a reference point. From here, the direction was determined by spinning a bottle or a pen and the *nth* interval between structures was identified using the second last digit of a bank note. The reference household was not included in the survey. In case of household refusal, the next household was approached. In District Malir, where we had a line listing of all households, a simple random sampling was done. In both areas, all household members were eligible to participate irrespective of their infection status. Approval from the household and written informed consent or assent from individual participants was obtained.

All team members underwent training in the use of PPE, hand hygiene and safe transportation of biological samples. A 3-mL sample of blood from infants and 5-mL from older participants, was collected by a trained phlebotomist and transported to the Infectious Diseases Research Lab for centrifugation, serum separation and storage at −20^0^C. Information was collected from all the participants on age, gender, occupation and household size, along with details of travel history and exposure to COVID-19 patients. Reported comorbidities, presence of symptoms, history of hospitalization we also recorded. An occupational history was also inquired from individual participants regarding working from home and otherwise. For symptoms, the clinical history recorded details of the presence of fever, respiratory symptoms such as sore throat, shortness of breath and chest pain in previous two months (Supplemental Appendix 3).

### Laboratory analysis

A commercial Elecsys® Anti-SARS-CoV-2 immunoassay (Roche Diagnostics), targeting combined IgG and IgM against SARS-CoV-2 was performed at the Nutritional Research Laboratory (NRL) at Aga Khan University. The manufacturer reported a specificity greater than 99.8% and sensitivity of 100% for individuals with a positive PCR test at least two weeks prior, and 88.1% sensitivity for those 7-13 days after a PCR-positive test (20).

## STATISTICAL ANALYSIS

### Sample Size

The sample size for each phase of the survey was calculated to be 500 participants for each site. This allowed for an estimation of an age-adjusted prevalence in the range of 20-30 % at 95% confidence level, with a precision of ± 5% and a design effect of 1.5 for household level clustering.

### Data Management

Data entry was performed in duplicate on an SQL database (Structured Query Language Database) (21) and was thoroughly checked for completeness and consistency. Continuous variables such as age and household size were reported as mean and standard deviation (SD). Categorical variables like gender, occupation and symptoms were reported as frequencies and percentages. Participants reporting fever or respiratory symptoms in the last two months were categorized as symptomatic and presented as proportions. Age was also categorized.

### Estimation of overall, age and gender stratified seroprevalence

Age and gender-stratified seroprevalence estimates were computed for each district and each phase independently, using a Bayesian hierarchical regression model. This approach, described in detail in the Supplementary Material, accounts for uncertainty due to finite lab validation data, (22) and produces estimates using typical choices of uninformative or weakly informative prior distributions (23, 24).

Given the correlation among household seropositivity values, model also took into account factors such as total number of members in a household and adjusted for test accuracy by modeling directly on the lab validation data reported by the test manufacturer (20). Seroprevalence estimates by age and gender were then post stratified to adjust for the demographic makeup of the respective districts.

Estimates are expressed as posterior means and 95% equal-tailed credible intervals based on 20,000 samples from a Bayesian posterior distribution. All calculations were performed in Software R and samples from posterior distributions were obtained using Stan (25).

### Household Conditional Risk of Infection (CRI) and Infection Fatality Rate analysis

**Conditional Risk of Infection** (CRI) is the probability that an individual is infected, conditioned on a household member being infected (26). CRI was calculated and presented as a fraction where the numerator is the total number of ordered pairs among infected individuals in the same household and whose denominator is the total number of ordered pairs in the same household in which the first individual in the pair is infected. A 95% confidence interval was estimated via bootstrap for each area by resampling households with replacement.

**Age specific - Infection Fatality Rate (IFR):** This measure estimates the prevalence of infection (including both asymptomatic and mildly symptomatic infections). The cumulative number of deaths due to COVID-19 infection were calculated till 14 days after sero-survey dissemination. This number was then divided by the figure obtained from multiplying the relevant population of an area with the adjusted estimate of seroprevalence (27).

## Results

### Participant Flow

A total of 3005 participants were enrolled across three phases from District East and District Malir (Figure 1 Panels, A, B and C). There were high refusal rates in both areas at the household level, 68%, 43% and 61 % in district East and 44%, 42% and 8% in district Malir, in phases 1, 2 and 3 respectively. The geographical location of households which volunteered to participate in the survey is presented in the supplemental figure 2 in appendix. Amongst the households who consented to participate, individual participation rate was 82.3% (1000 out of 1215 eligible household members) in phase 1, 76.5% (1004 out of 1312 eligible household members) in phase 2 and 80% (1001 out of 1243 eligible household members) in phase 3. Table 2 describes the baseline demographic and clinical characteristics of the enrolled participants.

**Figure 1 Panel A:**
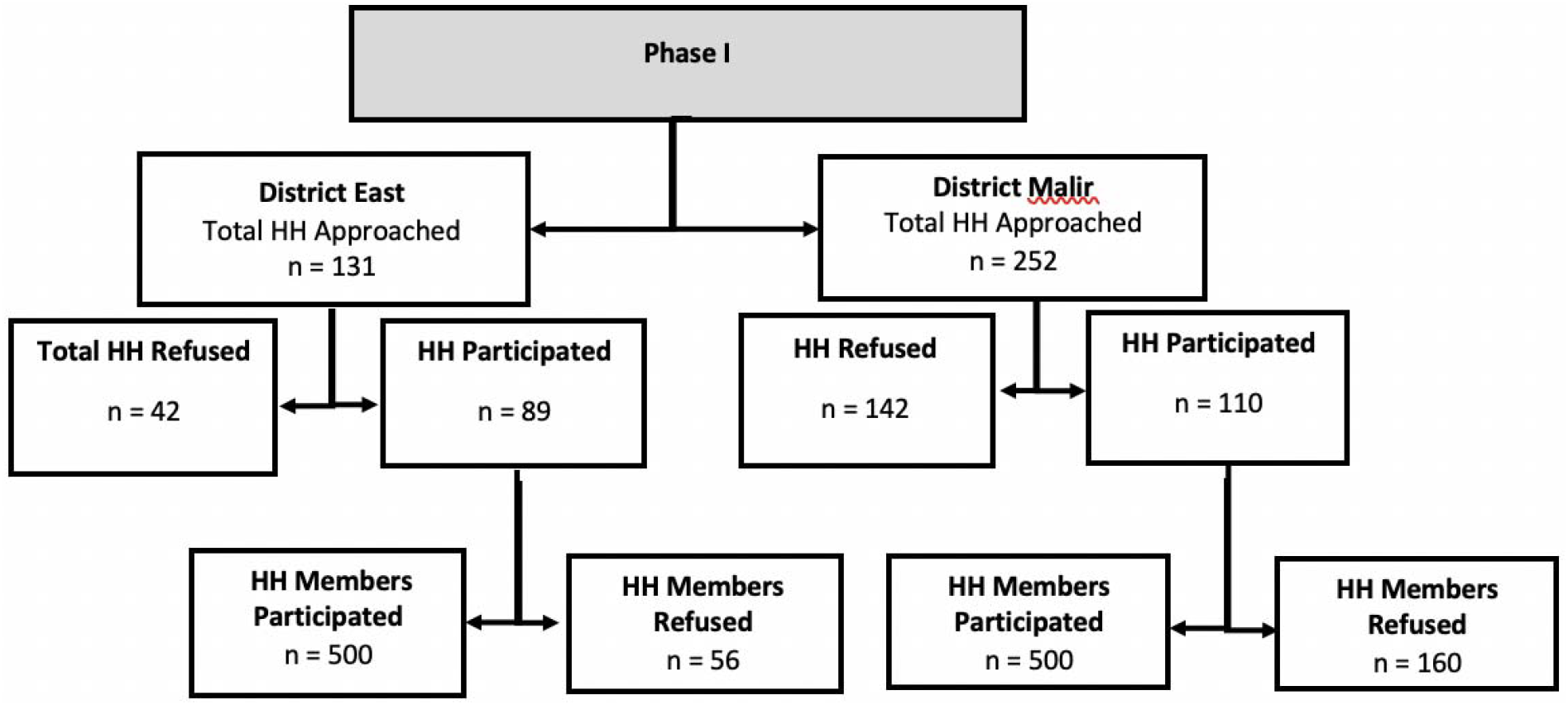
Flow chart of participants in Phase 1 of study

**Figure 1 Panel B.**
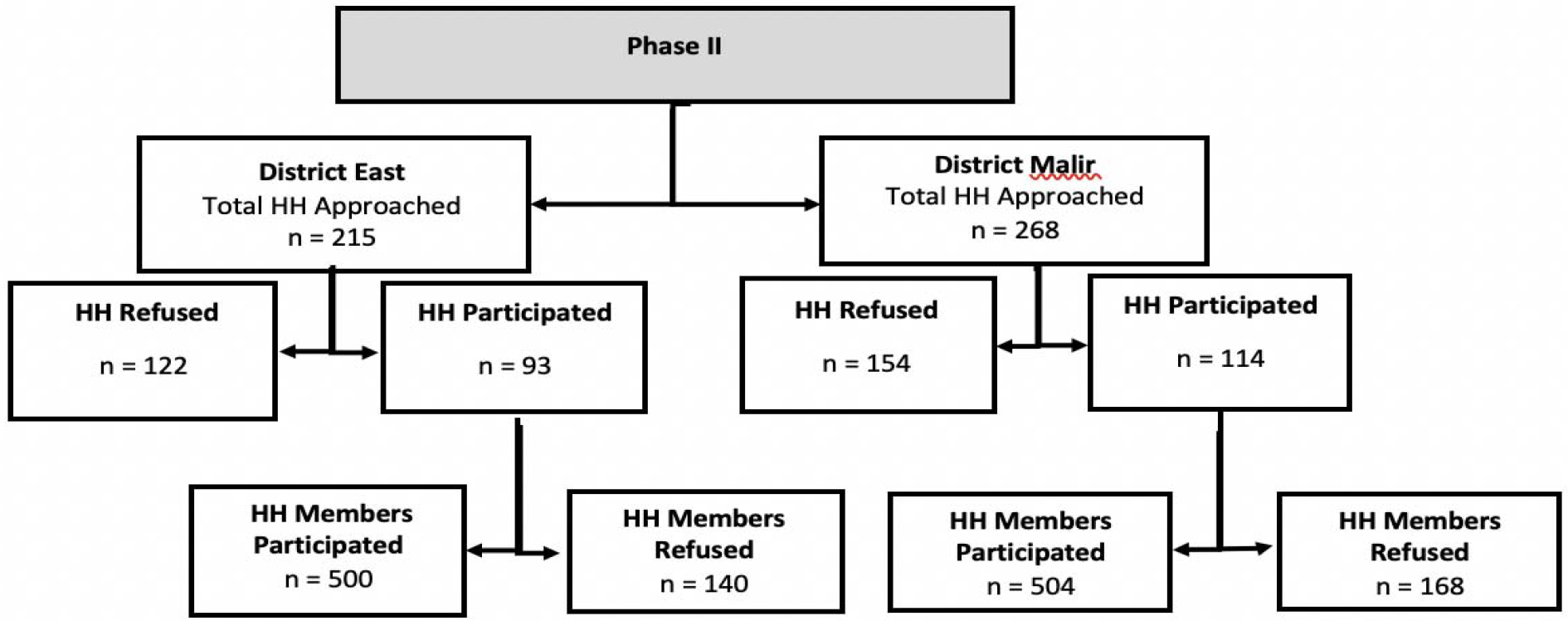
Flow chart of participants in Phase 2 of study.

**Figure 1 Panel C.**
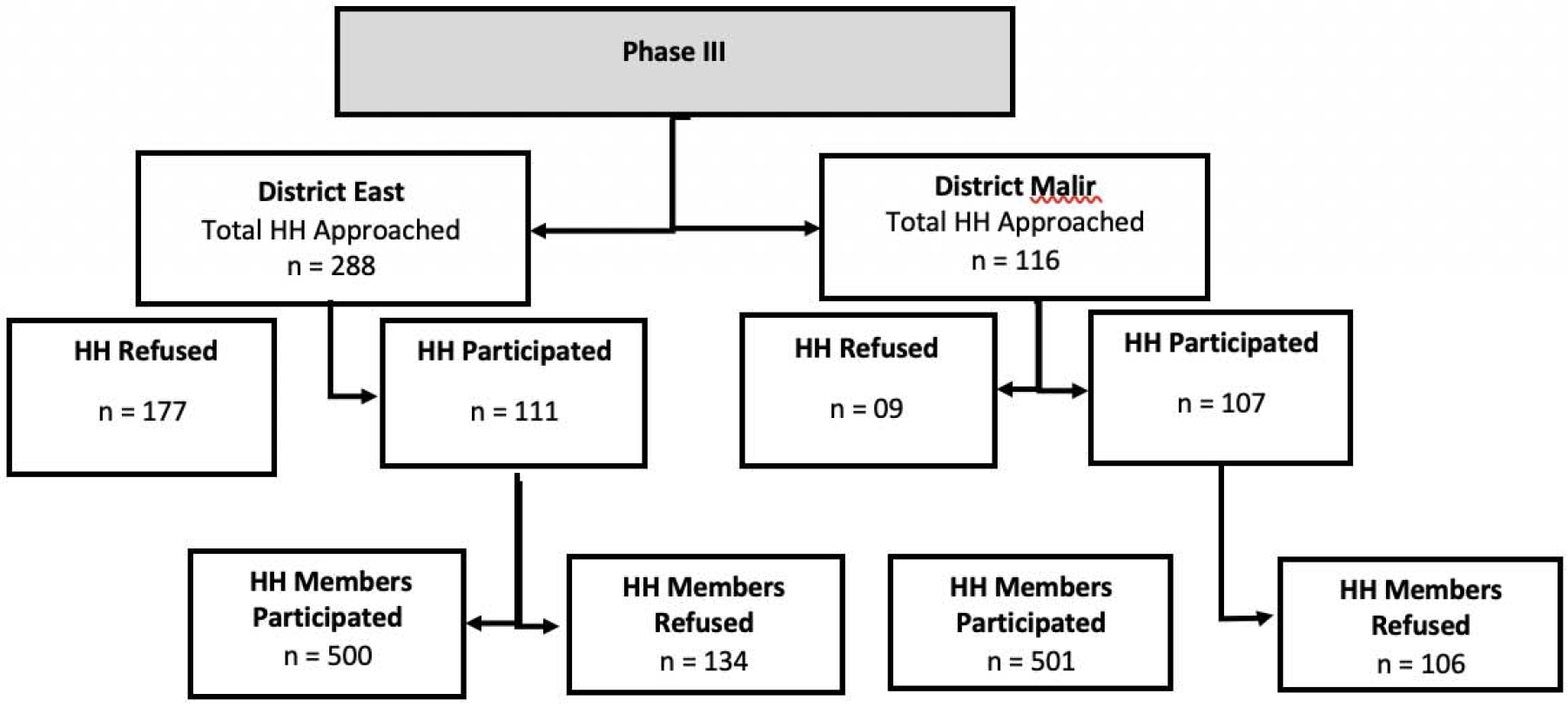
Flow chart of participants in Phase 3 of study.

### Seropositivity

In Phase 1 of the study, only 2 of 500 samples tested positive in District East, while none of the 500 participants tested positive in District Malir. In Phase 2 of the study, 100 of 500 samples (20.0%) tested positive in District East and 64 of 504 samples (12.7%) tested positive in District Malir. In phase 3 of the study, 119 of 500 samples (24 %) tested positive in District East and 79 of 501 samples (16%) tested positive in District Malir.

### Adjusted Seroprevalence

In Phase 1, post-stratified seroprevalence was estimated to be 0.4% (95% CI 0%-1.3%) in District East and 0.2% (95% CI 0%-0.7%) in District Malir. In Phase 2, it was estimated to be 15.1% (95% CI 9.4%-21.7%) in District East, and 8.7% (95% CI 5.1%-13.1%) in District Malir. Lastly, in Phase 3, the rate was documented to be 21.5% (95% CI 15.6%-28%) in District East, and 12.8 % (95% CI 8.3%-17.7%) in District Malir.

Both districts showed marked and significant increase in seroprevalence between phases sequentially with a sharp rise between Phase 1 and 2 and a smaller rise between phase 2 and 3. Overall reporting of symptoms was documented to be 9.4 %. Of the total 364 participants who tested positive, 27 (7.4 %) gave a history of fever or respiratory symptoms or both in the last 2 months (Supplementary Table 1).

### Conditional Risk of Infection and Infection Fatality Rates

To measure whether individuals in the same household were more likely to have similar sero-status, we computed the conditional risk of infection (CRI) for phases 2 and 3. CRI estimates were 0.41 (95% CI 0.28-0.52) and 0.38 (95% 0.27-0.52) in District East and 0.31 (95% CI 0.16-0.4) and 0.33 (95% 0.12-0.47) in District Malir in phases 2 and 3 respectively. In parallel, the age specific Infection Fatality Rate (IFR), a measurement calculated based on the cumulative total of infected persons (based on seroprevalence) and number of deaths (obtained from local government surveillance data), was calculated to be 1.66% in phase 1, 0.37% in phase 2 and 0.26%% in phase 3.

The rise in seroprevalence in district East corresponded with the epidemiology as ascertained through daily case reporting in the district as well as the four study sampling sites. (Supplementary Figure 3). Seropositivity rates were indistinguishable between male and female within each district as well as between age groups (Figure 2).

**Figure 2-A:**
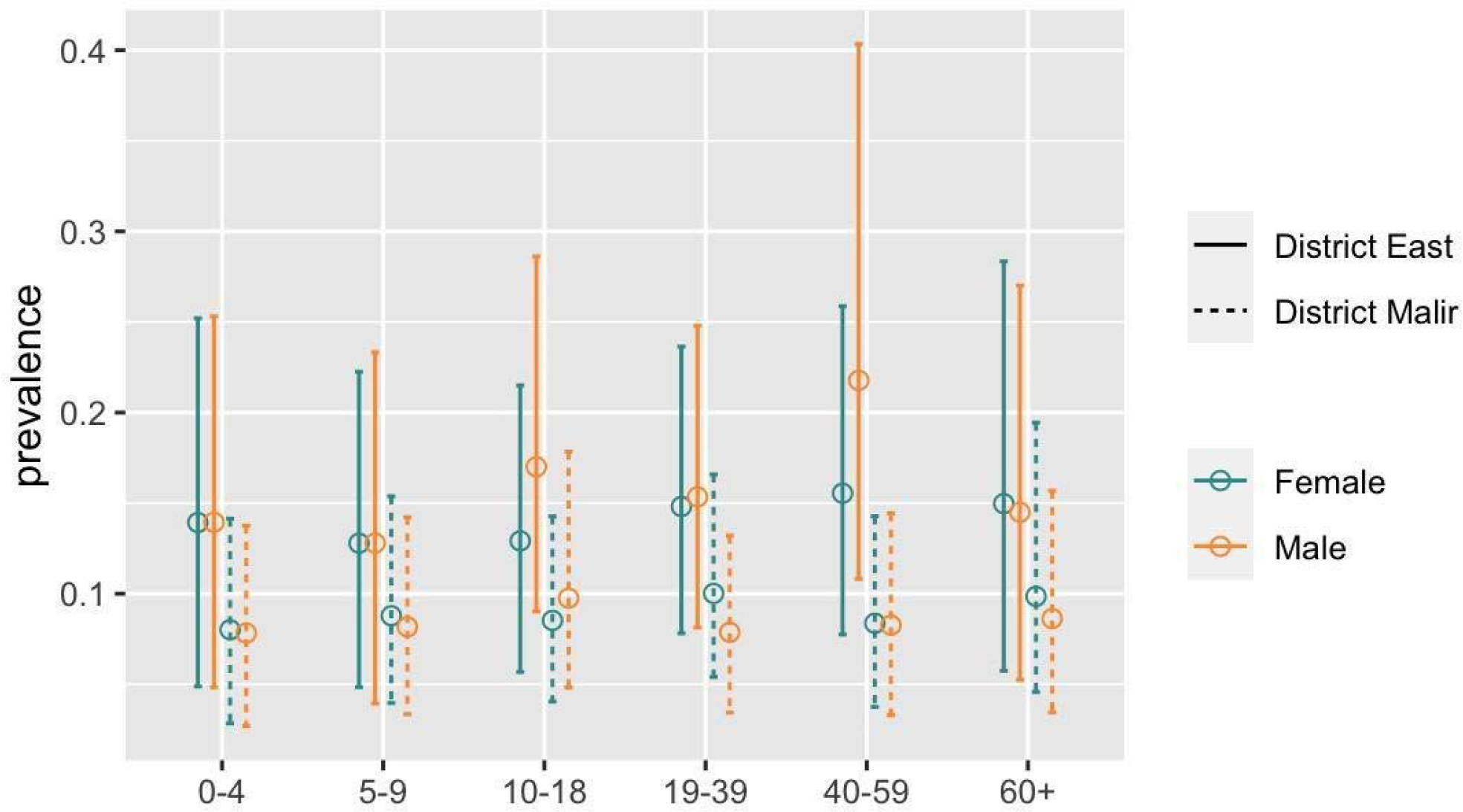
Age and Gender based prevalence of COVID-19 infection in District East and District Malir for Phase 2.

**Figure 2-B:**
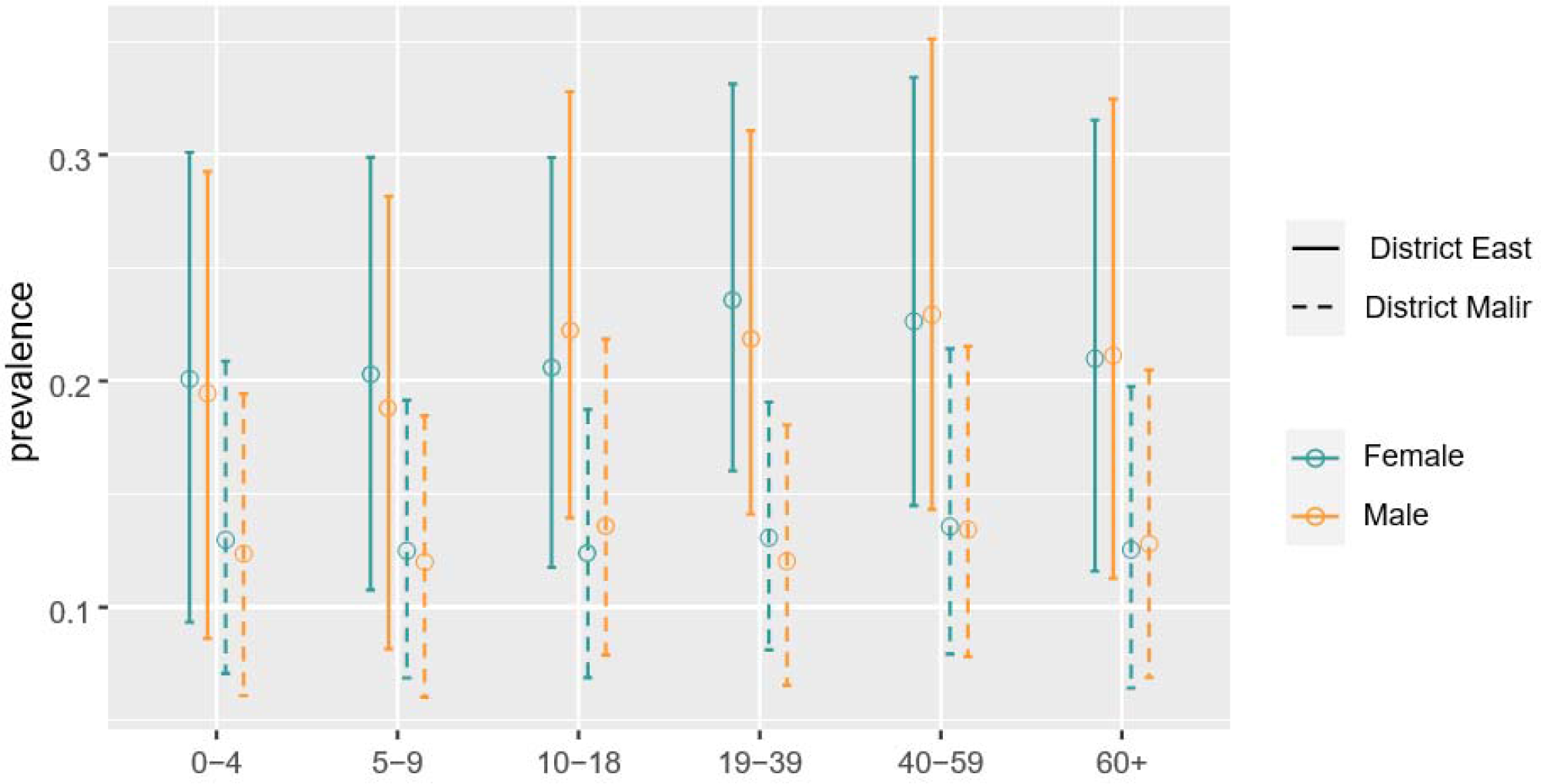
Age and Gender based prevalence of COVID-19 infection in District East and District Malir for phase 3

## Discussion

To our knowledge, this is the first published population level seroprevalence study in Pakistan, which provides baseline rates for comparison. Preliminary reports of seroprevalence studies from other metropolitan areas like Mumbai and Pune in India have highlighted high rates of seroprevalence, up to 55% (28), while a survey carried out in the Guilan province of Iran showed a prevalence of 33% at the peak of the pandemic (29, 30). In contrast, the three serial serosurveys conducted in the same populations with similar methodology in Karachi indicated an early increase, followed by a plateau in seroprevalence rates between the months of July and August, as demonstrated by Supplementary Figure 3. A number of reasons can explain the seroprevalence trends observed in our study. The first phase of the study was done early in the pandemic, 3 weeks after a provincial lockdown was declared while the second phase was conducted after lockdown measures were eased in anticipation of the religious festivities observed during Eid in Pakistan. Although individual associations cannot be discerned, effectiveness of social mitigation measures could explain the smaller rise in phase three. The survey did not identify any difference in seroprevalence between males and females or any age-specific trends in infection rates across different age categories. Prevalence appeared to increase with age and was documented to be consistently high between 19-39 years of age and 40-59 years of age in both males and females. This is consistent with age related seropositivity patterns in the literature (31). Our survey found a large number of asymptomatic sero-positive individuals. Only 3 out of 10 reported any respiratory symptoms, with or without fever. Consistent with our findings, the proportion of asymptomatic infections was reported to be much lower, 27.7% (95% CI, 16.4%□42.7%) in a meta-analysis (32). However, higher rates of asymptomatic infection have been reported in some parts of the world like India. According to WHO and the Indian Council of Medical Research (ICMR), India, the number of asymptomatic cases appears to be about 80% compared to the 20% that are symptomatic (33). Lower reporting of symptoms in Pakistan may be attributable to innate fears of disclosure of disease positivity and lack of awareness about the high transmissibility of the disease in the general public in the early stages of the pandemic.

The elevation in seroprevalence even in an area of presumably low transmission indicates that seroprevalence studies may serve as efficacious tools to determine spread of infection in populations where a large majority of the people are asymptomatic. Monitoring the general populations through serial sero-surveys can detect resurgence, especially when lockdown measures are eased and enable policy makers to devise strategies for containment of the disease. Heterogeneity between low- and high-income neighborhoods is likely and has been suggested previously (34-36). This cannot be conclusively deduced from our study where a different sampling strategy was used in the two neighborhoods. Census data for households in order to conduct simple random sampling was available only for District Malir, while systematic random sampling was used in District East around a reference household, reporting a case in the last 2 months. This may overestimate absolute seroprevalence in District East but enabled temporal comparisons of seroprevalence within the districts independently.

Our results also confirm that close contact within households is linked to a high probability of being infected and should be an important consideration in COVID-19 transmission (37). Our study indicated that the probability of an individual to acquire an infection in the presence of another infected household member, as measured by conditional risk of infection (CRI), was high between 35% - 40%. An unpublished review used a secondary attack rate, which is a more reliable indicator of intra-household transmission and reported it to be lower, i.e., 18.8% (95% CI 15.4%-22.2%) (38). CRI can hence, function as a substitute in situations where comprehensive surveillance efforts and disease notification strategy are absent and where secondary attack rates are difficult to estimate.

Our study indicated that the age specific IFR was higher in phase 1 (1.66%) and then decreased in phases 2 and 3 to 0.37% and 0.26% respectively. This may be attributed to early detection and improved management of symptomatic disease. A systematic review and meta-analysis has conceded that overall IFR could be 0.68% (0.53%-0.82%) but with heterogeneity from either the studies contributing to the review or due to various factors like age and presence of co-morbidities in the population (39). Population demographics can affect estimates of population-weighted IFR, with the lowest IFRs recorded for countries with younger populations, for instance Kenya (0.09%; 95% credible interval, 0.08–0.10%) and Pakistan ((0.16%; 95% credible interval, 0.14–0.19%) (40).

The strength of our study is that it filled the gaps in the baseline knowledge of seropositivity at a population-level, that was absent in most sero-surveys. The serial nature of our study, along with a two-month inter-survey interval allowed for a sequential evaluation of changes in sero-prevalence. About one third or more of our sample also included children less than 18 years of age, focusing on an understudied age group in the pandemic. Adaptation of the UNITY protocol also allowed for pooling of our results with other reports in future.

Our study had several limitations. The geographical area of the study was limited to only two neighborhoods and the sampling strategy differed, not allowing for comparisons between the areas. We also had high rates of household level refusal which can be attributable to the fact that the study was conducted in the midst of a pandemic when sentiments of fear and stigma were at their highest. Due to a limited supply chain of testing kits in Pakistan, we did not do an in-house validation on local samples, however this was compensated by modeling directly on the data reported by the manufacturer.

## Conclusion

A non-uniform increase has been documented in seroprevalence to SARS-CoV-2 in areas where disease transmission is reportedly low. The overall infection fatality rate has correspondingly decreased. Most sero-positive cases are reported to be asymptomatic and majority of the population is seronegative. However, there is a high probability of an individual to be infected following exposure within the same household, irrespective of symptoms. Enhanced surveillance efforts for COVID-19 are required on a national and provincial level, especially in low-transmission sites in order to determine the far-reaching consequences of the pandemic and the risk of household transmission in tightly knit neighborhoods in urban LMIC settings.

**Table 1.** General characteristics of the study participants

| Characteristics | Districts |  |  |  |  |  |
| --- | --- | --- | --- | --- | --- | --- |
|  | District East |  |  | District Malir |  |  |
|  | Phase 1 | Phase 2 | Phase 3 | Phase 1 | Phase 2 | Phase 3 |
|  | (n= 500) | (n= 500) | (n= 500) | (n= 500) | (n= 504) | (n= 501) |
| Sex, male n (%) | 256 (51.2%) | 225 (45.0%) | 211(42.2%) | 207 (41.4%) | 199 (39.5%) | 206(41.1%) |
| Age in years , mean(SD) | 26.2 (17.9) | 25.9(16.7) | 27.1(17.7) | 28.5 (17.9) | 24.32 (16.7) | 26(16.7) |
| Age, categories, n (%) |  |  |  |  |  |  |
| 0-4 year | 35 (7.0%) | 22 (4.4%) | 31(6.2%) | 26 (5.2%) | 33 (6.6%) | 29(5.8%) |
| 5-9 year | 57 (11.4%) | 54 (10.8%) | 56(11.2%) | 52 (10.4%) | 74 (14.7%) | 75(15.0%) |
| 10-18 year | 107 (21.4%) | 139 (27.8%) | 91(18.2%) | 86 (17.2%) | 120 (23.9%) | 87(17.4%) |
| 19-39 year | 185 (37.0%) | 170 (34.0%) | 206(41.2%) | 203 (40.6%) | 186 (37.0%) | 195(38.9%) |
| 40-59 year | 83 (16.6%) | 97 (19.4%) | 84(16.8%) | 103 (20.6%) | 65 (12.9%) | 90(18.0%) |
| 60 and above | 33 (6.6%) | 18 (3.6%) | 32(6.4%) | 30 (6.0%) | 25 (5.0%) | 25(5.0%) |
| Household size, mean (SD) | 6.1(4.17) | 6.6(3.6) | 5.7(2.3) | 6(2.8) | 5.89(3.2) | 5.6(2.6) |
| Working outside home, n (%) | 155 (31.0%) | 145 (29.0%) | 107(21.4%) | 143 (28.6%) | 124 (24.6%) | 116(23.2) |
| Comorbid, n (%) |  |  |  |  |  |  |
| None | 453 (92.3%) | 463 (93.0%) | 471(94.2%) | 473 (94.6%) | 479 (95.0%) | 482(96.2%) |
| Diabetes | 20 (4.0%) | 14 (2.8%) | 8(1.6%) | 6 (1.2%) | 10 (2.0%) | 5(1.0%) |
| Hypertension | 14 (2.8%) | 13 (2.6%) | 16(3.2%) | 16 (3.2%) | 11 (2.2%) | 14(2.8%) |
| Asthma or Allergy | 3 (0.6%) | 3 (0.6%) | 3(0.6%) | 1 (0.2%) | 0 (0.0%) | 0(0.0%) |
| Chronic Hepatitis | 2 (0.4%) | 3 (0.6%) | 1(0.2%) | 3 (0.6%) | 3 (0.6%) | 0(0.0%) |
| Chronic Heart Disease | 0 (0.0%) | 3 (0.6%) | 1(0.2%) | 1 (0.2%) | 1 (0.2%) | 1(0.2%) |
| Asymptomatic | 435 (87.0%) | 453 (90.6%) | 471(94.2%) | 472 (94.4%) | 487 (96.6%) | 473(94.4%) |
| Sought care | 9 (1.8%) | 9 (1.8%) | 2(0.4%) | 3 (0.6%) | 4 (0.8%) | 0(0.0%) |
| Hospitalization | 1 (0.2%) | 5 (1.0%) | 0(0%) | 1 (0.2%) | 0 (0.0%) | 0(0.0%) |
| History of Travel | 2 (0.4%) | 11 (2.2%) | 7(1.4%) | 8 (1.6%) | 1 (0.2%) | 4(0.8%) |
| Contact with suspected or confirmed COVID-19 case | 1 (0.2%) | 1 (0.2%) | 2(0.4%) | 0 (0.0%) | 1 (0.2%) | 1(0.2%) |

## Data Availability

Data will be made available on request

## Acknowledgments

The authors would like to acknowledge all the data collectors, phlebotomists and laboratory personnel who made this happen in the most difficult of circumstances.

---

Figure 1 Panel A. Flow chart of participants in the phase 1 of study

Figure 1 Panel B. Flow chart of participants in the phase two of study

Figure 2: Prevalence estimates by age and gender based on the data from the second survey. The circle represents the posterior mean seroprevalence and the bar represents the 95% equal-tailed credible interval. Posterior mean estimates for District East are consistently greater than those for District Malir, although there is significant overlap in the credible intervals for all age and gender subpopulations. No consistent patterns exist between the prevalence rates for males and females.

