## Supplemental Material for "Serial population-based sero-surveys for COVID-19 in low and high transmission neighborhoods of urban Pakistan"

**Supplementary Table 1**. Distribution of participants on the basis of presence of symptoms and test reactivity

|  | Non-reactive  N=2641 | Reactive  N=364 | Total  N=3005 |
| --- | --- | --- | --- |
| Symptomatic |  |  |  |
| Fever  Fever & respiratory symptoms*  Only respiratory symptoms* | 46 (1.7)  39 (1.5)  104 (3.9) | 10 (3)  10 (3)  7 (1.9) | 56 (1.9)  49 (1.6)  111 (3.7) |
| Asymptomatic or Other Symptoms | 2452 (92.8) | 337(92.6) | 2789 (92.8) |
| Fatigue  Muscle Ache (Myalgia) | 10 (0.4)  21 (0.9) | 0 (0.0)  1 (0.3) | 10 (0.4)  22 (0.8) |
| Headache  Nausea/Vomiting  Abdominal Pain  Diarrhea  No Symptoms at All | 39 (1.6)  0 (0.0)  3 (0.1)  2 (0.1)  2388(94%) | 3 (0.9)  1 (0.3)  1 (0.3)  0 (0.0)  333 (98.8) | 42 (1.5)  1 (0.0)  4 (0.1)  2 (0.1)  2721(98.1) |

*includes cough, sore throat, shortness of breath, chest pain, wheezing, runny nose

| **Variables** | | **Phase 2** | | **Phase 3** | |
| --- | --- | --- | --- | --- | --- |
| **Gender** | **Age** | **District East** | **District Malir** | **District East** | **District Malir** |
| Female | 0-4 | 0.139 (0.04, 0.25) | 0.080 (0.02, 0.14) | 0.201(0.093, 0.301) | 0.13(0.071, 0.209) |
| Female | 5-9 | 0.128 (0.04, 0.22) | 0.088 (0.04, 0.15) | 0.203(0.108, 0.299) | 0.125(0.069, 0.192) |
| Female | 10-18 | 0.130 (0.05, 0.21) | 0.086 (0.04, 0.14) | 0.206(0.118, 0.299) | 0.124(0.069, 0.187) |
| Female | 19-39 | 0.149 (0.07, 0.23) | 0.101 (0.05, 0.16) | 0.236 (0.16, 0.331) | 0.131(0.081, 0.191) |
| Female | 40-59 | 0.155 (0.07, 0.26) | 0.084 (0.03, 0.14) | 0.226(0.145, 0.334) | 0.136(0.079, 0.214) |
| Female | 60+ | 0.149 (0.05, 0.27) | 0.099 (0.04, 0.19) | 0.21(0.116, 0.315) | 0.125(0.064, 0.198) |
| Male | 0-4 | 0.139 (0.04, 0.25) | 0.079 (0.02, 0.14) | 0.195(0.086, 0.292) | 0.124(0.061, 0.194) |
| Male | 5-9 | 0.128 (0.03, 0.23) | 0.082 (0.03, 0.14) | 0.188(0.082, 0.281) | 0.12(0.06, 0.185) |
| Male | 10-18 | 0.170 (0.09, 0.28) | 0.098 (0.04, 0.17) | 0.222(0.139, 0.328) | 0.136(0.079, 0.219) |
| Male | 19-39 | 0.153 (0.08, 0.24) | 0.079 (0.03, 0.13) | 0.219(0.141, 0.311) | 0.12(0.065, 0.181) |
| Male | 40-59 | 0.217 (0.10, 0.40) | 0.083 (0.03, 0.14) | 0.229(0.143, 0.351) | 0.134(0.078, 0.215) |
| Male | 60+ | 0.145 (0.05, 0.26) | 0.087 (0.03, 0.15) | 0.211(0.113, 0.325) | 0.128(0.069, 0.205) |

**Supplementary** **Table 2**: Sero-prevalence posterior mean and 95% equal-tailed credible interval estimates based on the second and third survey for both districts.

**Supplementary Figure 1**. Study area and total population.

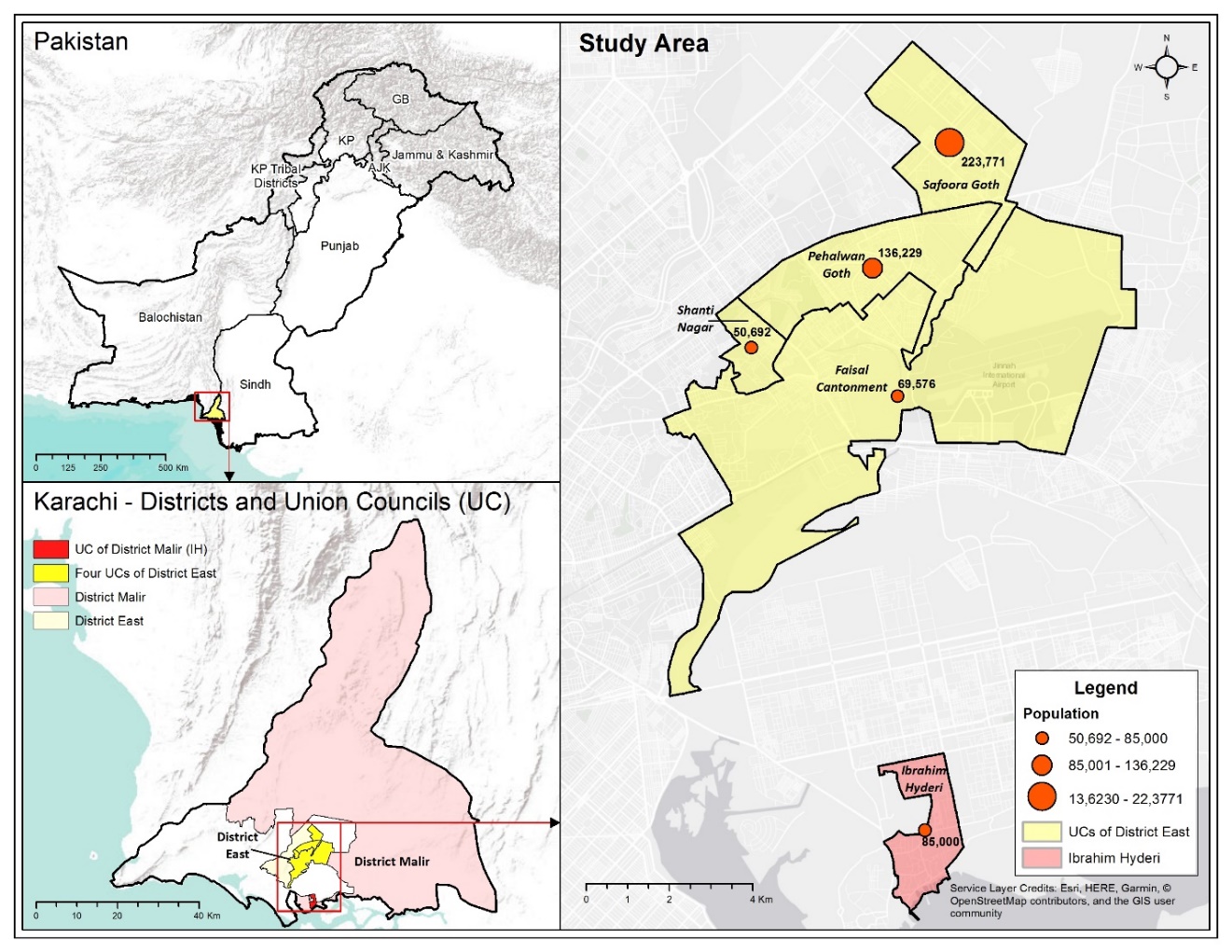

**Supplementary Figure 2**: Map showing GIS location of enrolled and refused households in the study sites (selected Union Councils of District Malir and District East, Karachi).

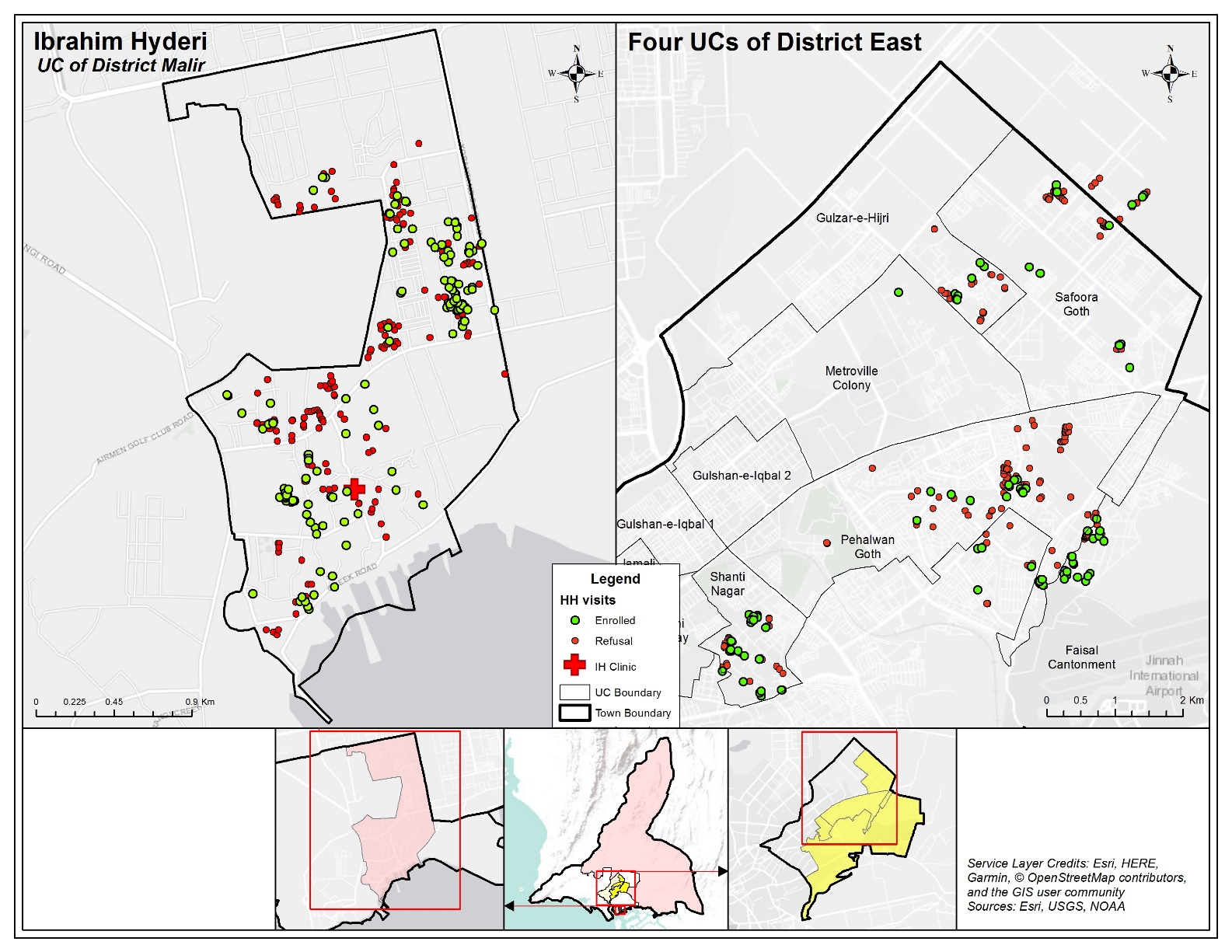

**Supplementary Figure 3.** Comparison of COVID-19 positive cases trend from the District East and study sampling sites with an estimation of seroprevalence in Phases 1, 2 and 3.

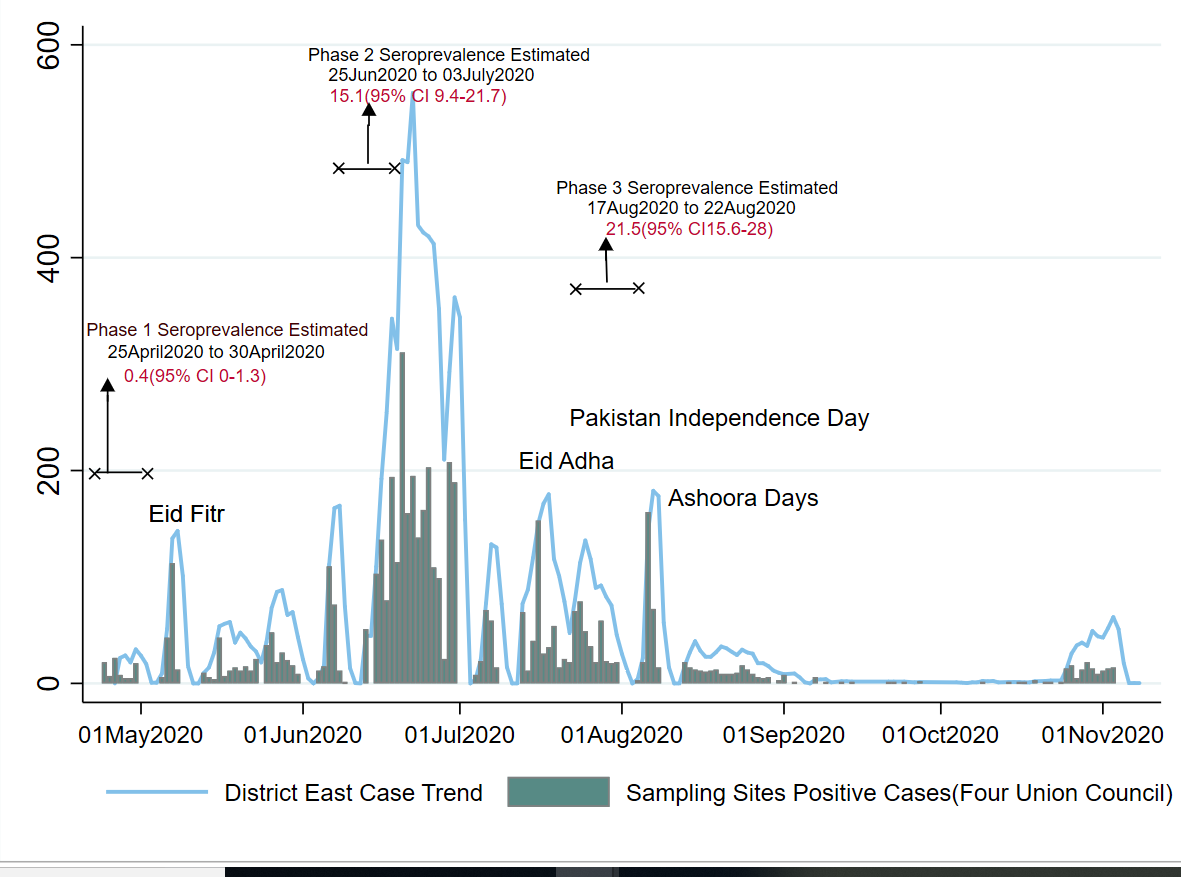
